# Stigmatizing Language Detection in Opioid Use Disorder Patient-Directed Discharge Clinical Documentation: A Privacy-Preserving Analysis Using a Locally Deployed Large Language Model

**DOI:** 10.64898/2026.05.29.26354402

**Authors:** Joseph A. Izzo, Adrienne M. McIntyre, Jacklyn Nguyen, Dennis Bashaw, Caprice A. Torrance, Jennifer Foster

## Abstract

**Objective:** Stigmatizing language in the electronic health record (EHR) has been associated with adverse patient experience in substance use disorder care, including opioid use disorder (OUD). This study evaluated a privacy-preserving, locally-deployed large language model as a method to detect stigmatizing language documentation in OUD patients with patient-directed discharge (PDD).

**Methods:** A retrospective cohort study of 477 inpatient admissions from the MIMIC-IV database with a diagnosis of opioid use disorder were classified using a locally deployed Gemma-4-31b-it-bf16 model and predefined 140 term lexicon to identify stigmatizing language in clinical documentation.

**Results:** Analysis of clinical documentation showed stigmatizing language was present in 84.1% (190/226) in the PDD cohort vs 62.2% (156/251) in the non-PDD cohort, with an unadjusted odds ratio of 3.21 (95% CI 2.07–4.98; p < 0.0001). After adjustment for age, sex, insurance status, marital status, and race, PDD discharge remained an independent predictor of stigmatizing documentation (aOR 2.24, 95% CI 1.40–3.59; p < 0.0001). Further analysis of stigma intensity showed higher stigmatizing markers in the PDD cohort vs the non-PDD cohort (2.85 ± 2.39 vs 2.02 ± 2.44; p < 0.0001).

**Discussion and Conclusion:** Stigmatizing language is detected with increased frequency and prevalence in clinical documentation of OUD patients that initiate PDD compared to those that adhere to standard discharge processes. A locally deployed large language model (LLM) offers a scalable, privacy-preserving method to audit clinical documentation for stigmatizing language.

## Introduction

Stigmatizing language in the electronic health record (EHR) has been associated with adverse patient experiences leading to patient-directed discharge (PDD) in substance use disorder (SUD) care, including opioid use disorder (OUD).[1] Stigmatizing language, consisting of spoken or written words that communicate unintended meanings and perpetuate socially constructed power dynamics resulting in bias.[2] Stigmatizing language can include words or phrasing that leads to discrimination against a patient based on their “health conditions” or socio-economic conditions.[3] The National Institute of Drug Abuse recommends using “person with substance use disorder” instead of addict, “person in active use” instead of junkie, or “person with opioid addiction” instead of user.[4] When a clinician reads a note laden with biased phrasing, it can lead them to view the patient as a problem rather than a person seeking help, which may directly compromise the quality of care. Studies have shown that the treatment of patients with OUD by resident physicians was changed by stigmatized versus non-stigmatized language found in the EHR, demonstrating that stigma is transmitted and can affect the patient before they are seen by the physician.[5, 6, 7] Terminology choice in clinical documentation may influence patient trust, engagement, and continuity of care.

Traditional approaches to auditing documentation language have relied on manual chart review or keyword-based natural language processing (NLP).[8] These methods are either not scalable or fail to capture context, particularly in cases involving negation, subject attribution (e.g., patient vs. family history), or shorthand clinical phrasing. Large language models (LLMs) offer a fundamentally different approach by enabling contextual interpretations of clinical texts rather than relying on surface-level lexical features.[8] Recent work by Sethi et al. demonstrated the feasibility of LLM-based stigma detection in ICU progress notes.[9] We build upon this work by examining whether patient-directed discharge is independently associated with stigmatizing documentation, using a privacy-preserving, locally deployed pipeline intended for implementation in resource-limited settings.

Use of foundation or cloud-based models raises legitimate concerns regarding exposure of sensitive patient data.[10] Recent advances in model optimization, including quantization and efficient local inference frameworks, have enabled high-performing LLMs to run on a single workstation GPU rather than large clusters of high-end accelerators, allowing for contextual language analysis while lowering infrastructure costs and preserving patient privacy through local data retention.

Patient-directed discharge, previously referred to as discharge against medical advice (AMA), represents a clinically meaningful outcome in OUD care. According to Garneau et al., persons with OUD are 10 to 20 times more likely to engage in PDD and PDD has been shown to “increase mortality, readmission, and overall costs for health systems”.[11] Prior work by Leon, et al. demonstrated that stigmatization in healthcare settings negatively affects patient experience and engagement in care.[1] Documentation language is one mechanism through which stigma is conveyed and reinforced, yet difference between stigmatizing and non-stigmatizing language associated within a diagnostically homogenous population remains incompletely characterized.[5, 6, 7]

Here we present a privacy-preserving, secure, locally-deployed large language model pipeline to audit OUD-related clinical documentation, comparing predefined linguistic markers considered stigmatizing between PDD and non-PDD cohorts.

## Methods

### Data and study setting

We conducted a retrospective cohort study of 477 inpatient admissions from the Medical Information Mart for Intensive Care (MIMIC-IV, Version 3.1) database with a recorded diagnosis of opioid use disorder (Figure 1).[12] The MIMIC-IV database contains de-identified inpatient admissions from Beth Israel Deaconess Medical Center (BIDMC) spanning 2008 to 2019 and is compliant with HIPAA de-identification standards.[8] A predefined 14-term lexicon was used to identify candidate sentences potentially containing stigmatizing language related to substance use documentation. Lexicon terms were matched using substring matching, such that a term like ‘addict’ would also capture ‘addiction’, ‘addicted’, and related forms. Lexicon terms were derived from established substance use stigma literature, including National Institute on Drug Abuse (NIDA) “Words Matter” guidance and prior published studies evaluating stigmatizing terminology in clinical documentation.[4, 11] Documentation sentences were then classified using a locally deployed Gemma-4-31b-it-bf16 model (https://huggingface.co/mlx-community/gemma-4-31b-it-bf16) with rule-constrained prompts targeting behavioral stigma, selected for final analysis based on improved contextual precision over initial piloting with Llama 3.3-70B. Sentences with documented administrative PDD terminology, (e.g.: against medical advice, eloped, signed forms, leaving, advice), were excluded via keyword filtering. To assess the construct validity of the classification framework, a random sample of 100 sentences (n=100) underwent manual review and comparison with model classification outputs. The adjudication framework generated structured rationales for each classification, enabling traceability of model decisions and facilitating targeted review of ambiguous or discordant cases. Stigma prevalence was compared using Fisher’s Exact test; marker frequency was compared using the Mann-Whitney U test. Logistic regression adjusted for age, race, sex, insurance, and marital status. This research utilized secondary de-identified information from a publicly-available dataset and was deemed exempt, not meeting the definition of human research, by the Institutional Review Board at San Joaquin General Hospital. Access to MIMIC-IV data was limited to study personnel who completed required CITI training and established appropriate credentialing and Data Use Agreement requirements through PhysioNet.

**Figure 1.**
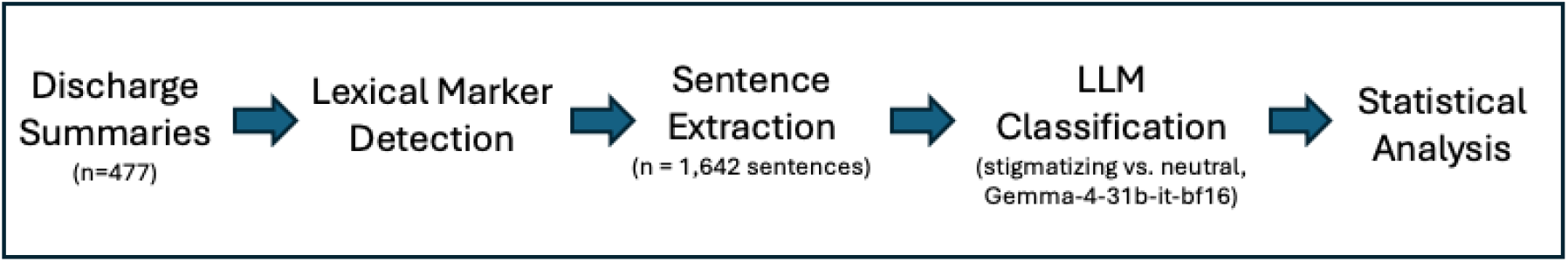
Analytical pipeline for detection of stigmatizing language in discharge summaries. Candidate sentences were identified via lexicon-based screening, followed by contextual adjudication using a locally deployed large language model (Gemma-4-31b-it-bf16), and statistical comparison between patient-directed discharge (PDD) and control admissions.

All code was executed on a local Apple Mac Studio M3 Ultra. The MIMIC-IV database was hosted locally via PostgreSQL v14.18. Inference was performed locally using Python 3.12 with the MLX framework (0.31.2).[13] Models were obtained via HuggingFace (https://huggingface.co). Statistical analysis was performed using Numpy (version 2.4.2),[14] SciPy (version 1.17.1),[15] and pandas (version 3.0.). Large language model assistance (Claude Sonnet 4.6, Anthropic) was used during development of SQL and Python scripts. Any AI-generated code was reviewed by authors (J.A.I) with domain expertise in SQL and Python for accuracy, potential hallucinations, and analytical integrity. No AI-generated text was incorporated into the manuscript without author review and revision.

### Outcome variables

A total of 477 inpatient admissions associated with opioid use disorder were analyzed, including 226 admissions resulting in patient-directed discharge (PDD) and 251 non-PDD admissions. Following lexicon-based screening, contextual adjudication using a locally deployed Gemma 4 model classified 1,156 of 1,642, 70.4%, candidate sentences as stigmatizing. After exclusion of 7 sentences containing administrative PDD terminology (e.g. “AMA”, “Against medical advice”, “eloped”), 1,149 sentences remained classified as stigmatizing. Among candidate sentences identified by the lexicon, 29.6% were adjudicated as neutral clinical language, reflecting the role of contextual evaluation in reducing false positive classifications.

## Results

Descriptive statistics of stigmatizing language in the clinical documentation identified prevalence and frequency; 84.07% (190/226) of discharge summaries in the PDD cohort compared with 62.15% (156/251) in the non-PDD cohort (Figure 2). This corresponded to an unadjusted odds ratio of 3.21 (95% CI 2.07–4.98; p < 0.0001, Fisher’s exact test). After adjustment for age, sex, insurance status, marital status, and race, PDD discharge remained an independent predictor of stigmatizing documentation (aOR 2.24, 95% CI 1.40–3.59; p < 0.0001) (Figure 3 and Table 1).

**Table 1.** Baseline demographic characteristics of patients with opioid use disorder by discharge group. PDD = patient-directed discharge. P-values derived from Mann–Whitney U test for continuous variables, Fisher’s exact test for binary variables, and chi-square test for race.

| Variable | PDD (n=226) | Control (n=251) | p-value |
| --- | --- | --- | --- |
| Age, mean ± SD | 40.0 ± 11.0 | 48.1 ± 14.5 | |
| Age, median (IQR) | 38.0 (31.2–48.0) | 49.0 (36.5–60.0) | <0.0001 |
| Female sex, n (%) | 82 (36.3%) | 93 (37.1%) | 0.9242 |
| Race, n (%) |  |  | 0.7605 |
| White | 158 (69.9%) | 180 (71.7%) |  |
| Black | 27 (11.9%) | 28 (11.2%) |  |
| Hispanic | 12 (5.3%) | 17 (6.8%) |  |
| Other | 29 (12.8%) | 26 (10.4%) |  |
| Medicaid Insurance, n (%) | 160 (70.8%) | 132 (52.6%) | <0.0001 |
| Married, n (%) | 18 (8.0%) | 41 (16.3%) | 0.0077 |

**Figure 2.**
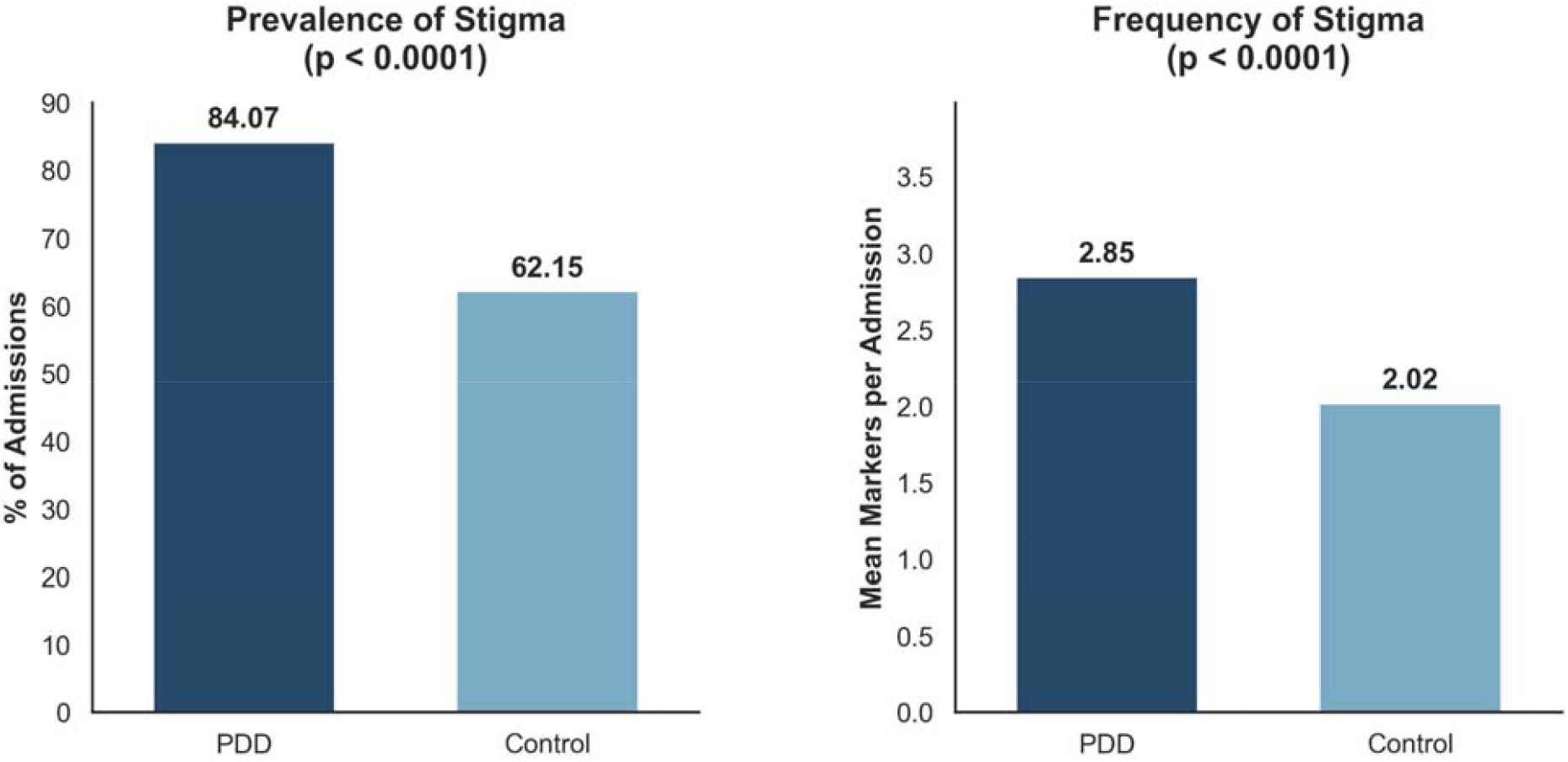
Prevalence (left) and frequency (right) of stigmatizing language in discharge summaries among patients with opioid use disorder. PDD = patient-directed discharge. Fisher’s exact test and Mann–Whitney U test, respectively (p < 0.0001 for both).

**Figure 3.**
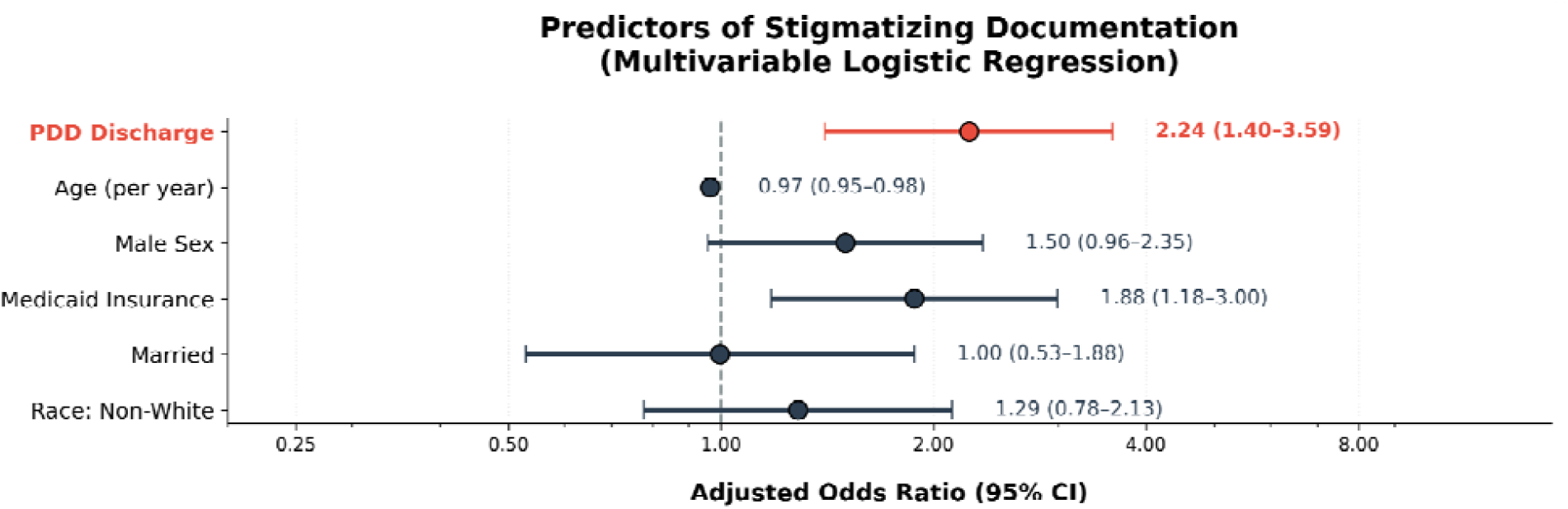
Multivariable logistic regression forest plot showing adjusted odds ratios (aOR) for predictors of stigmatizing documentation in discharge summaries. PDD discharge was the primary predictor of interest (red). Horizontal lines represent 95% confidence intervals. Reference line at aOR = 1.0.

Analysis of stigma intensity demonstrated a higher mean number of stigmatizing markers per admission in the PDD cohort compared with the non-PDD cohort (2.85 ± 2.39 vs 2.02 ± 2.44) (Figure 4). This difference was statistically significant using the Mann–Whitney U test (p < 0.0001). Secondary parametric analysis yielded consistent results.

**Figure 4.**
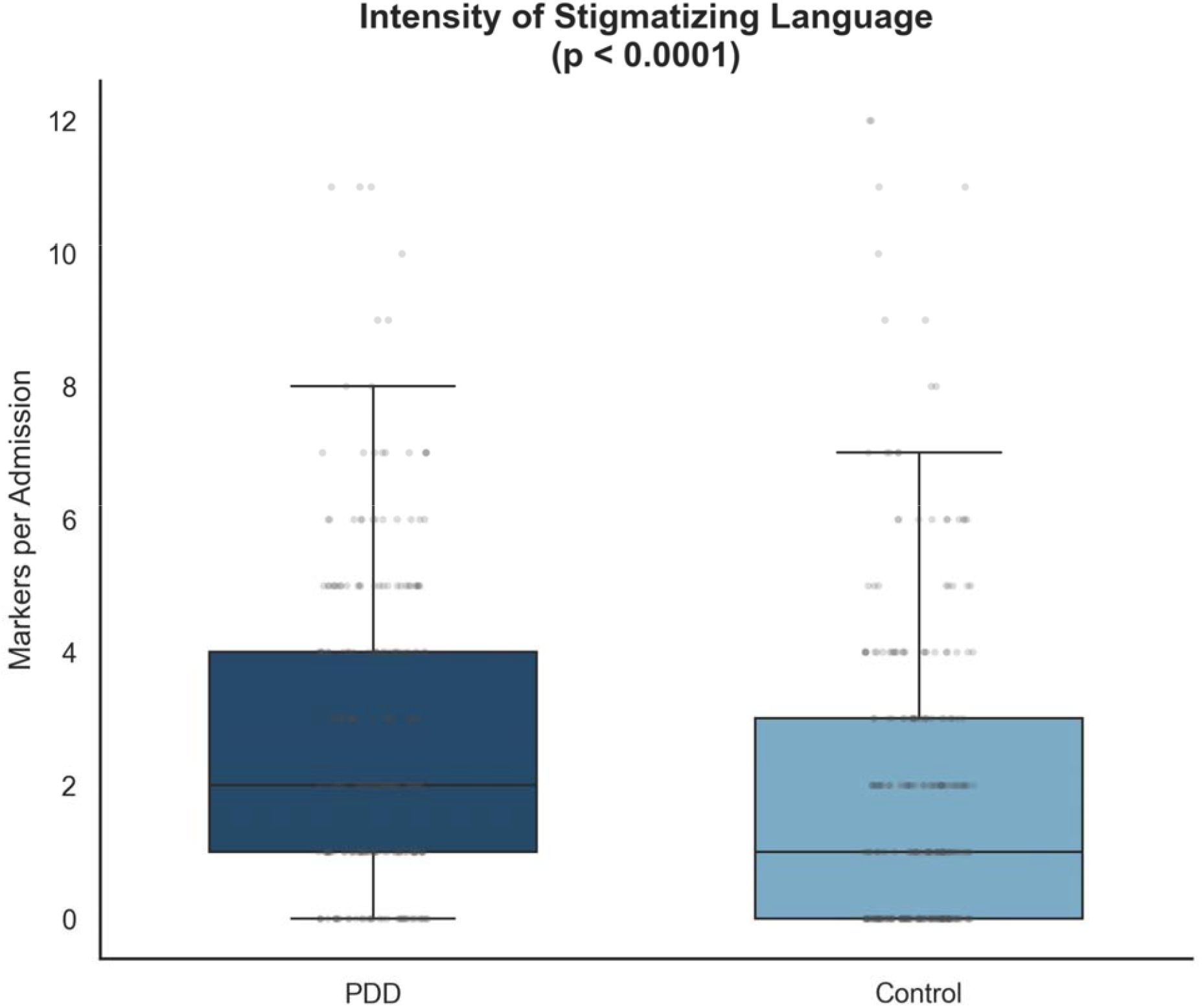
Distribution of stigmatizing language intensity per admission among patients with opioid use disorder by discharge group. Boxes represent interquartile range; horizontal line indicates median. Outliers shown as individual points. PDD = patient-directed discharge. Mann–Whitney U test (p < 0.0001).

Word-level analysis demonstrated that legacy substance use descriptors such as “abuse” and “Intravenous drug use (IVDU)” accounted for the highest frequency of stigmatizing classifications. Behavioral descriptors such as “refused,” which implies a level of “difficulty” or lack of compliance with treatment recommendations, were also frequently classified as stigmatizing, with contextual adjudication identifying stigmatizing usage in 83.8% of occurrences (Figure 5 and Table 2). In contrast, terms matching “addict” and “frequent” demonstrated lower rates of stigmatizing classification, reflecting context-dependent usage. Qualitative review also identified clinically relevant contextual language patterns not detectable through lexicon matching alone, including documentation framing patient behavior in pejorative or characterological terms, supporting the utility of contextual LLM-based adjudication.

**Table 2.** Prevalence of stigmatizing language by category and most frequent terms in discharge summaries among patients with opioid use disorder by discharge group. Values represent the number of admissions containing at least one instance of the term (%) [total instances]. PDD = patient-directed discharge. P-values derived from Fisher’s exact test.

| Category/Term | PDD | Control | p-value |
| --- | --- | --- | --- |
| **Language category** |  |  |  |
| Any stigmatizing language | 190 (84.1%) | 156 (62.2%) | <0.0001 |
| Lexical (legacy labels) | 172 (76.1%) | 147 (58.6%) | < 0.0001 |
| Behavioral descriptors | 71 (31.4%) | 39 (15.5%) | < 0.0001 |
| Mixed | 2 (0.9%) | 2 (0.8%) | 1.0000 |
| **Most frequent terms** |  |  |  |
| Abuse | 119 (52.7%) [274] | 107 (42.6%) [260] | 0.0346 |
| IVDU | 99 (43.8%) [204] | 67 (26.7%) [142] | 0.0001 |
| refused | 59 (26.1%) [103] | 30 (12.0%) [55] | < 0.0001 |
| addict | 22 (9.7%) [24] | 8 (3.2%) [11] | 0.0041 |
| clean | 12 (5.3%) [15] | 11 (4.4%) [13] | 0.6734 |
| frequent | 8 (3.5%) [9] | 6 (2.4%) [8] | 0.5893 |

**Figure 5.**
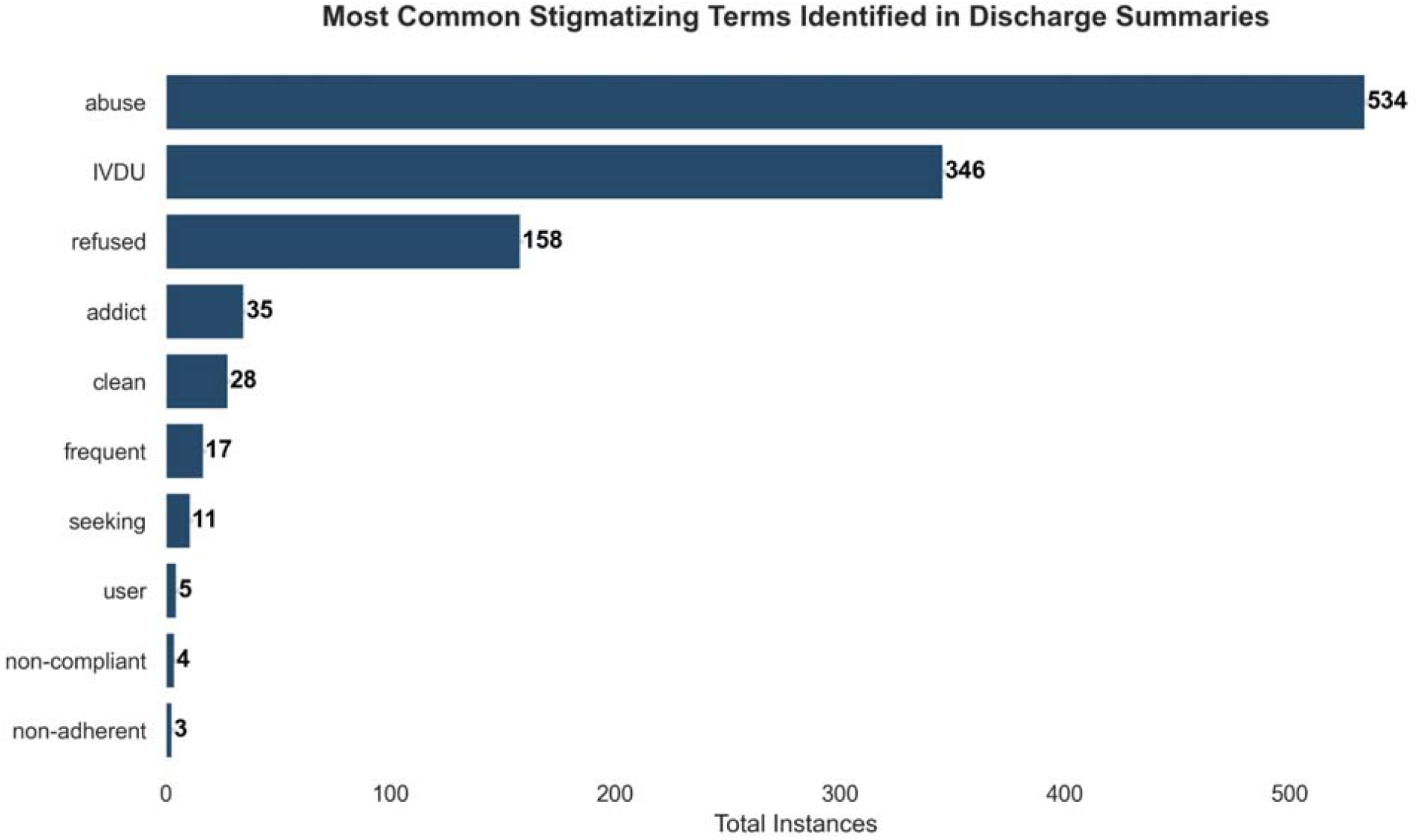
Most frequently identified stigmatizing terms across all discharge summaries (N=477). Values represent total instances identified following LLM adjudication and administrative term exclusion. Terms were matched using substring matching.

## Discussion

Differences in documentation between stigmatizing and non-stigmatizing patient notes associated with PDD appear to be driven by narrative framing rather than diagnostic content. These findings suggest that clinical documentation reflects not only clinical status but also the framing of patient behavior. Patient groups with inherently higher risk behaviors, like OUD, may encourage the use of language and terminology intended to defer blame to the patient when opting for care that is deemed misaligned with clinical recommendations. This inherently shifts the clinical note documentation from focus on the patient to focus on protection of the clinician.

In modern medicine, the EHR often reaches the physician before the patient does. Kelly and Westerhoff found that referring to a patient as “a substance abuser” vs. “having a substance use disorder” triggers different responses, punishment vs. treatment.[6] The chart assumes the patient’s identity leading to continual stigmatization and prejudice and lower quality of care and poor outcomes.

The hybrid NLP-LLM approach allows scalable auditing while preserving patient privacy. The locally developed model functions as a rule-constrained contextual adjudicator rather than an autonomous classifier, aligning with governance and data protection considerations.

### Strengths and limitations

This study strongly demonstrates the ability to use a large language model to identify stigmatizing language in clinical documentation, reducing the need for a manual audit process. Our study had several limitations. The retrospective design and use of discharge summaries limit causal interpretation. Sample size was modest and consisted of patients and clinical documentation from one medical center - findings should be considered exploratory. The LLM was not formally evaluated for sensitivity, specificity, or recall. Differences in clinical documentation may reflect clinician perception of challenging encounters and may contribute to downstream patient experience, however, lack of outcome data beyond PDD limits the ability to link stigmatizing language documentation prevalence to patient harm or health inequities.

## Conclusion

Requirements for implicit bias training to address health inequities have provided education but there are limited ways to measure the impact of such training. Patient-directed discharge is associated with higher prevalence and frequency of stigmatizing language in documentation within an OUD cohort. A locally deployed LLM offers a scalable, privacy-preserving approach to auditing documented clinical language. This model for detecting stigmatizing language may offer a quantifiable way to assess stigmatizing language prevalence without the burden of manual chart review. Future applications of the LLM model may include prospective chart review and prevalence detection, rather than retrospective, to explore how a LLM model may be applied to quality and performance improvement methods within a healthcare organization.

## Data Availability

The data underlying this study are available through the MIMIC-IV database on PhysioNet to qualified researchers who complete the required training and data use agreements.

## Author’s Contributions

All authors contributed to the design, analysis, interpretation, drafting, or reviewing of this work. All authors approve of the final version of this work. CRediT contributor roles are as follows: J.A.I.: Conceptualization, data curation, formal analysis, methodology, project administration, resources, software, supervision, validation, visualization, writing-original draft, writing-review & editing. A.M.M.: Conceptualization, visualization, writing-original draft, writing-review & editing. J.N.: Writing-original draft, writing-review & editing. D.B.: Methodology, writing-original draft, writing-review & editing. C.A.T.: Resources, software, visualization, writing-review & editing. J.F.: Resources, software, visualization, writing-review & editing.

